# Oxidative stress and PARP activation in the lungs is an early event in COVID-19 pneumonia

**DOI:** 10.1101/2024.09.03.24312996

**Authors:** Péter Juhász, Péter Bohus, Adrienn Sipos, Nicola Curtin, Gábor Méhes, Péter Bai

**Author notes:** Correspondence can be also sent to: Peter Bai, PhD, DSc University of Debrecen, Faculty of Medicine, Department of Medical Chemistry, 4032 Debrecen, Egyetem tér 1., Hungary, Correspondence can be also sent to: Gábor Méhes, MD, PhD, DSc, University of Debrecen, Faculty of Medicine, Department of Pathology, 4032 Debrecen, Nagyerdei krt. 98., Hungary. joint first authorship.

## Abstract

Oxidative stress and poly(ADP-ribosyl)ation (PARylation) leads to tissue damage and inflammation in multiple lung diseases, likely in COVID-19. In a previous study we evidenced PARylation in multiple pulmonary cell types in patients who died of COVID-19, but not in patients who died of non-COVID-19 causes. We extended these observations in this retrospective immunohistochemical study by enlarging and stratifying the study population. We showed that pulmonary PARylation and oxidative stress peaked in the exudative and then decreased in the proliferative phase. Pulmonary oxidative stress and PARylation correlated with the serum markers of liver and kidney damage, oxygen transport, tissue hypoxia, lymphocytopenia, blood clotting and disseminated intravascular coagulation. Most correlations between PARylation and serum chemistry readouts were identified in the exudative phase. PARylation correlated with viral load and with the oxidative stress in the tissues, however, correlation between viral load and oxidative stress was marginal suggesting that oxidative stress and the presence of SARS-CoV-2 can independently induce PARylation. In males the time of hospitalization (time to death) was inversely correlated with pulmonary PARylation. Furthermore, males, died of COVID-19, were ∼15 years younger than females, however, there was no difference in pulmonary oxidative stress and PARylation between genders at death. Taken together, pulmonary PARylation and oxidative stress manifests early, in the exudative phase of COVID-19 and PARylation contributes to worse clinical outcome for males. These results suggest repurposing pharmacological PARP inhibitors for acute COVID-19 to counteract tissue damage.

## Introduction

Coronoavirus disease 2019 (COVID-19), caused by SARS-CoV-2 was the third leading cause of death in the United States in 2022 [1]. The etiology of COVID-19 disease is not fully established, nevertheless, oxidative tissue damage was evidenced in multiple organs [2-7] that likely has pathogenic significance.

PARP enzymes constitute a family of 17 members in humans [8-10], of which PARP1 and PARP2 are responsible for the bulk of cellular PARP activity (reviewed in [11]). PARP1, PARP2 and PARP3 can be activated by oxidative stress-evoked DNA damage [8-10]. When activated, PARPs cleave NAD+ into ADP-ribose and nicotinamide, with the ADP-ribose units being transferred onto acceptor proteins, generating poly(ADP-ribose) (PAR) that is a marker of PARP activation. This process is called PARylation. Cellular PARP activity is responsible for the conversion of a large fraction of cellular NAD+ to nicotinamide and ADP-ribose [12,13].

PARP activation is associated with multiple events related to COVID-19 pneumonia (listed in **Table 1** and reviewed in [14]), as oxidative stress-induced apoptotic or necrotic tissue damage [10], inflammation [15,16], acute respiratory distress syndrome [17], fibrosis [18-22], or mechanical lung injury [23-25]. PARP activation was demonstrated in the lung biopsies of patients who died of COVID-19 [7]. In this study we aimed to assess how PARP activity changes with the resolution of COVID-19 and whether PARP activation may contribute to differences in the outcome of the disease between sexes.

**Table 1.** Common features of low NAD+ levels, COVID-19 and PARP activation.

| **Predisposing factor for severe or fatal COVID-19** | **Relation to PARP activation** |
| --- | --- |
| Inflammatory response and cytokine storm [26]. | PARP1 and PARP2 are proinflammatory [15,16]. |
| Advanced age [27,28]. | PARP activity increases in aging [29-32]. |
| Metabolic diseases (type II diabetes, metabolic syndrome, and obesity) [33,34]. | PARP overactivation acts as a metabolic disruptor and contributes to obesity and diabetes [35,36]. |
| Mitochondrial damage and metabolic rearrangement [37-39] | PARP activation disrupts mitochondrial oxidation [40]. |
| Men are at higher risk [41-43]. | PARP activity is higher in men [44-46]. |
| Cardiovascular diseases [47,48]. | PARP activation contributes to cardiovascular diseases [49-52]. |

## Materials and methods

### Patients

The study was authorized by the local ethical board (6043/2022). Demographics of patient cohorts are in **Table 2**.

**Table 2.** Patient characteristics. For comparing the age between all and female lung and all non-lung pathology patients normality was checked using the Shapiro-Wilk test then one-way ANOVA test was applied and all groups were compared in all combinations; none of the comparisons proved to be significant. For comparing the age between male lung and all non-lung pathology patients normality was checked using the Shapiro-Wilk test then Kruskal-Wallis test was applied and all groups were compared in all combinations; none of the comparisons proved to be significant except of the comparison between males and females (p<0.001). For comparing age of between COVID-19 patients in the exudative vs. proliferative phase normality was checked using the Shapiro-Wilk test then a non-paired parametric t-test was applied; the difference was not significant. Abbreviations: AVG – average

| | Sex | COVID-19 stage | Number of patient | Age (years; AVG±SD) |
| --- | --- | --- | --- | --- |
| **COVID-19 patients** | All | | 28 (100%) | 68.8±12.4 |
| | Female | | 14 (50%) | 76.6±10.7 |
| | Male | | 14 (50%) | 59.8±7.9*** |
| | | Exudative | 17 (60.7%) | 71.0±14.3 |
| | | Proliferative | 11 (39.3%) | 65.3±8.1 |

We assessed all patients who died of COVID-19 at Szent Erzsébet Hospital (Sátoraljaújhely, Hungary) between 30^th^ November 2020 – 6^th^ February 2021. A part of this patient cohort was assessed already [7], in this study we complemented that study by assessing all patients who died of COVID-19 during the indicated period. We excluded all patients with nosocomial infections and one young female COVID-19 patient due to long-standing alcoholism (alcoholism induces PARP activity, e.g. [53]). A total of 40 patients were enrolled to the study, of which 28 fulfilled the enrollment criteria. COVID-19 patients had severe pneumonia involving both lungs (90-100% area inflamed). Patients were stratified as a function of the stage of the disease (exudative vs. proliferative) based on the histology findings that was verified by two independent pathologists; no ambiguous cases were identified.

### Medical records

The time of hospitalization was counted in days and each 24 hours that was begun was counted as s whole day (e.g. 36 hours of hospitalization counts as 2 days).

Interim blood tests (if performed) were reviewed and the results for 1) admission, 2) closest to/before death and 3) with the maximum/minimum of change in a parameter were identified. If one single blood test result was available that result was used as admission, death and maximum/minimum data.

### Immunohistochemistry

The description of the immunohistochemistry protocol is provided in the Supplementary materials.

### Statistical evaluation

GraphPad Prism v. 8.0.1 was used for statistics. Statistical tests are indicated in the figure or table captions.

## Results

### Oxidative stress and PARylation is stronger in the exudative versus the proliferative phase of COVID-19

Previously we showed that markers of oxidative stress and PARylation are present in the lung biopsies of patients who died of COVID-19. In this study we investigated all patients who died in the specified time window. Immunohistochemical sections of all eligible patients were scored for spike, 4HNE and PAR immunostaining intensity and the percent of 4HNE+ and PAR+ cells was determined and patients were stratified as a function of whether they died in the exudative or proliferative phase of COVID-19 [54]. All histological scores were higher in patients who died in the exudative versus those died in the proliferative phase (**Figure 1A**). Endothelial cells and pneumocytes were spike and PAR positive in all cases in both phases. Pneumocytes were 4HNE positive all cases in both phases. Endothelial cells were positive for 4HNE in most cases; the proportion of 4HNE negative cases was 29% in the exudative and 9% in the proliferative phase (**Figure 1B**).

**Figure 1.**
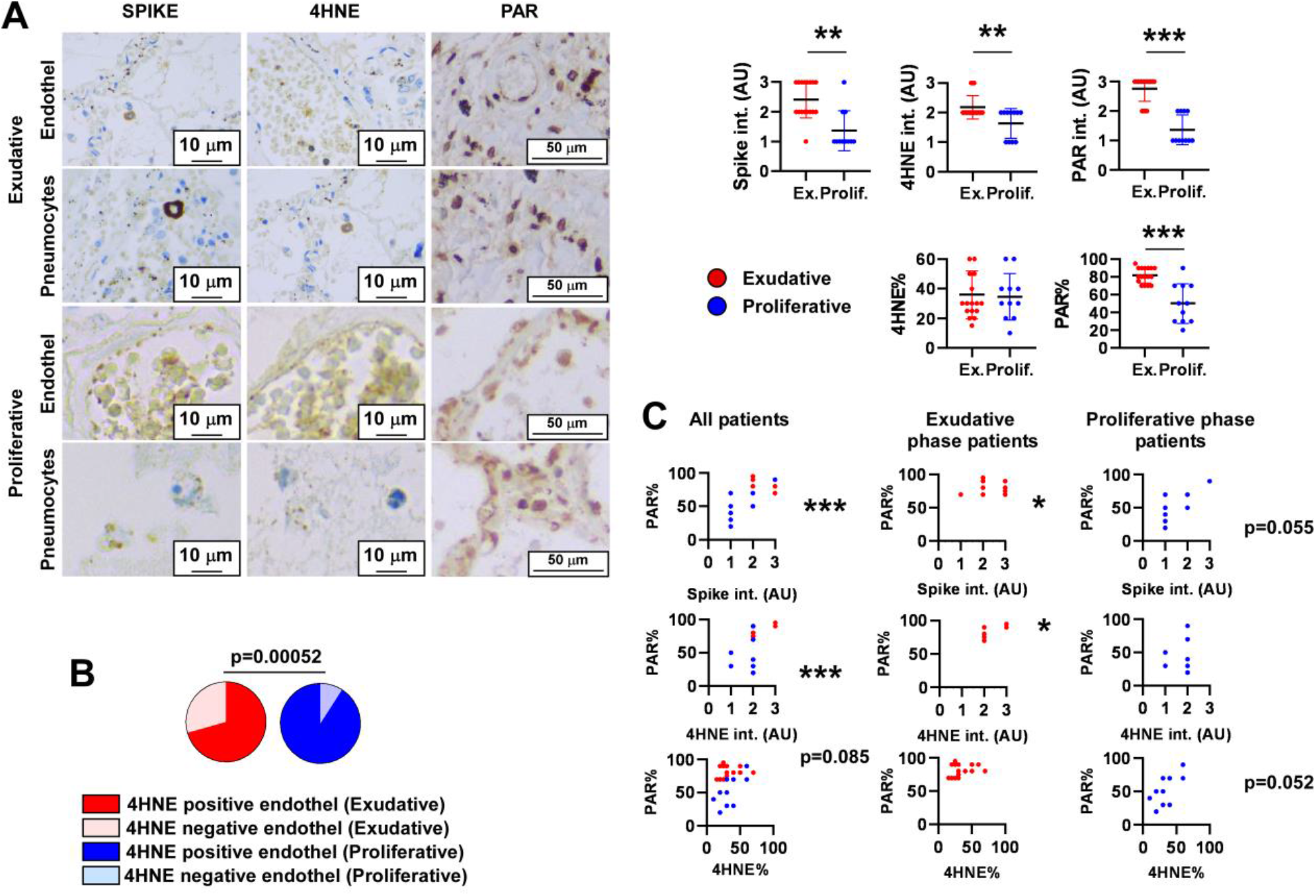
Oxidative stress and PARP activity is higher in the exudative than in the proliferative phase of COVID-19. **(A)**The indicated immunohistochemistry reactions were scored for the cases in the patient cohorts. Scale bar is 10 µm. Normality was checked using the Shapiro-Wilk test. In all cases Mann-Whitney test was applied to assess statistical significance. Average ± SD are depicted. ** or *** symbolize statistically significant difference between the indicated groups at p<0.01 or 0.001, respectively. **(B)**The proportions of the entohelial PAR positive and negative cases were plotted for patients died in the exudative and the proliferative phase of the disease. Statistical significance was assessed using the khi-square test. **(C)**Correlation between the indicated histology indices were calculated. Normality was checked using the Shapiro-Wilk test. The test applied, exact p and Rho values are listed in **Table S3**. * or *** symbolize statistically significant correlation between the indices marked on the axes at p<0.05 or .0.001, respectively.

PAR% correlated with spike intensity and 4HNE intensity in all patients, in exudative or in proliferative cases (**Figure 1C, Table S2**). However, there was only a borderline correlation between 4HNE indices and spike intensity (p=0.06), suggesting that the two PARylation inducers are likely unrelated (**Figure 1C, Table S2**).

These histology scores were correlated with serum parameters 1) when all patients were considered as one group and 2) patients in the exudative and in the proliferative phase were considered separately. 4HNE intensity or 4HNE% correlated positively with Na^+^ concentration, hemoglobin and ferritin levels, but negatively n with total Ca^2+^ concentration, red blood cell distribution volume, lymphocyte number and thrombin time (**Table 1**). PAR intensity or PAR% correlated positively with Na^+^, K^+^, Cl^-^ and Mg^2+^ ions, urea, direct bilirubin and ferritin levels, but negatively with total Ca^2+^ concentration, lymphocyte number and lymphocyte percent (**Table 3**). Correlations with PARylation and 4HNE implicate impact on kidney and liver function, oxygen transport and hypoxia, white blood cells and blood clotting or disseminated intravascular coagulation (DIC).

**Table 3.**
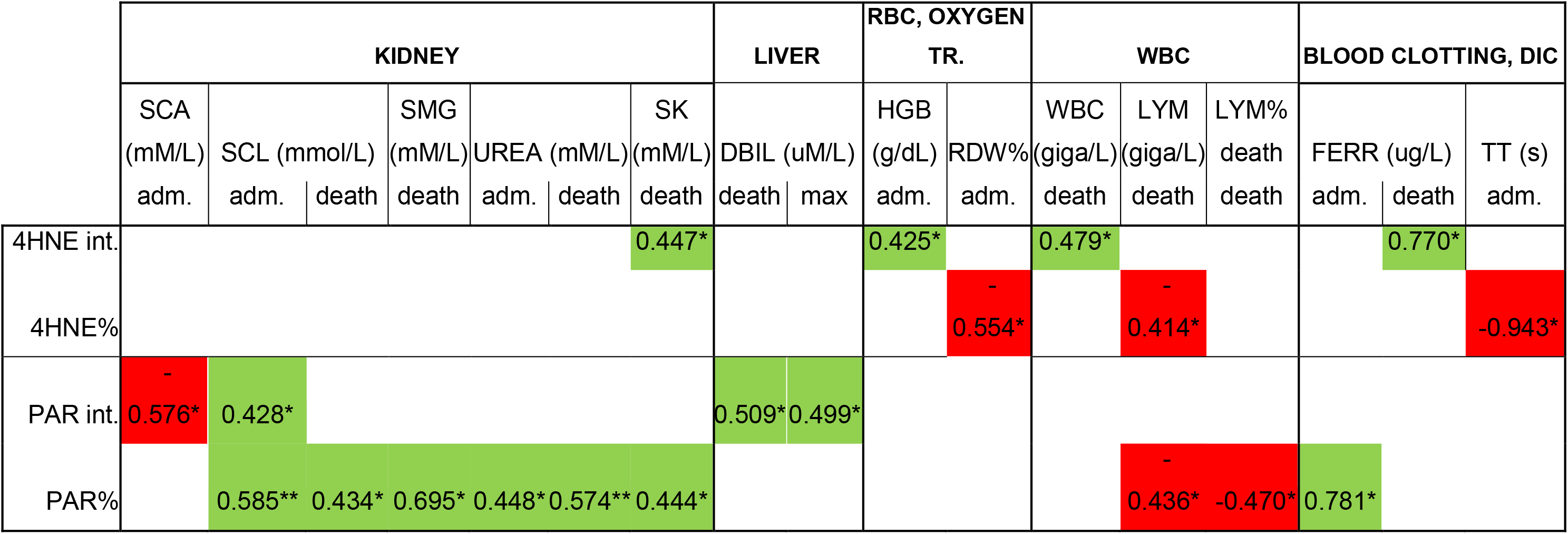
Pulmonary 4HNE and PAR scores correlate with serum chemistry and hemogram readouts. The serum chemistry and hemogram parameters of the COVID-19 patients were correlated with 4HNE intensity, 4HNE%, PAR intensity and PAR% using Spearman correlation. In the table significance is marked by asterisks and the numerical values represent the rho value. Green colors represents positive, while red color represent negative correlation. * or ** symbolize statistically significant correlation at p<0.05 or 0.01, respectively. Abbreviations: SK – serum potassium ion concentration, SCL -serum chloride ion concentration, SCA -serum total calcium ion concentration, SMG -serum magnesium ion concentration, DBIL – serum direct bilirubin concentration, HGB – hemoglobin concentration, RDW – red blood cell distribution, WBC – white blood cell count, LYM – lymphocyte count, LYM% -lymphocyte percent, FERR – serum ferritin, TT – thrombin time, RBC – red blood cell, DIC – disseminated intravascular coagulation, adm. – admission, int. -intensity.

In the exudative phase the indices of oxidative stress did not correlate with serum markers, while the indices of PARylation positively correlated with Na^+^ and urea concentrations, total and conjugated bilirubin concentration, gamma-glutamyl transferase and lactate dehydrogenase activity suggesting correlation with kidney and liver dysfunction and general tissue damage (**Table 4**). In the proliferative phase oxidative stress indices positively correlated with Na^+^ and Cl^-^ concentration and white blood cell count, furthermore, negatively correlated with K^+^ concentrations (**Table 4**). In the proliferative phase indices of PARylation positively correlated with Cl^-^ concentration and negatively with direct bilirubin, lymphocyte count and PCT% (**Table 4**).

**Table 4.**
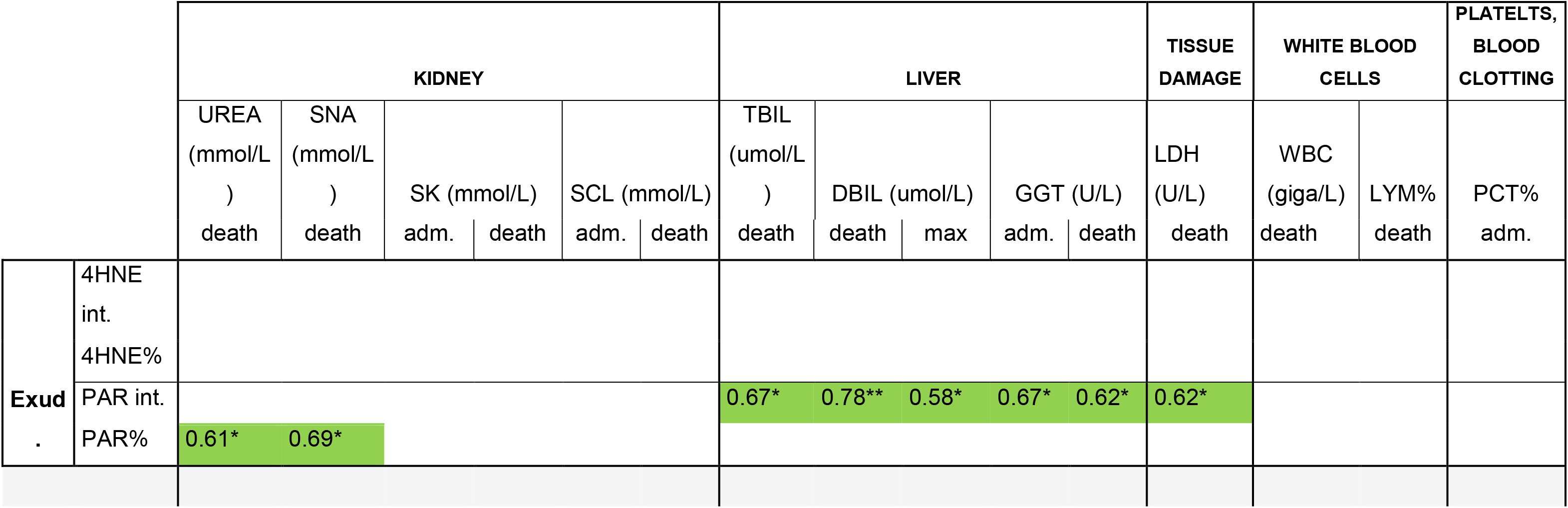
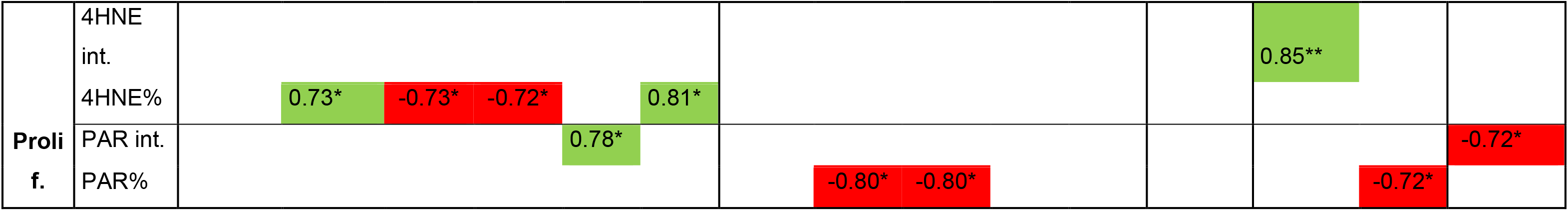
Pulmonary 4HNE and PAR scores have different correlation pattern with serum chemistry and hemogram readouts in the exudative and proliferative phase of COVID-19. The serum chemistry and hemogram parameters of the COVID-19 patients were correlated with 4HNE intensity, 4HNE%, PAR intensity and PAR% using Spearman correlation. In the table significance is marked by asterisks and the numerical values represent the rho value. . *, ** or *** symbolize statistically significant difference between the indicated groups at p<0.05, 0.01 or .0.001, respectively. Abbreviations: Exud. – exudative, Prolif. – proliferative, SNA – serum sodium ion, SK – serum potassium ion concentration, SCL -serum chloride ion concentration, TBIL – serum total bilirubin concentration, DBIL – serum direct bilirubin concentration, GGT – serum gamma-glutamyl transferase activity, int. – intensity, LDH – serum lactate dehydrogenase activity, WBC – white blood cell count, LYM% -lymphocyte percent, PCT% - plateletcrit, adm. – admission, DIC – disseminated intravascular coagulation.

### Oxidative stress and PARylation in the lungs supports negative outcomes in males

Mindful that PARylation levels are higher in males in non-COVID19 models [44-46] similar to the fatality rate of COVID-19 [41-43,55] we assessed sex-associated differences. Surprisingly, there was no difference between genders in histological indices, moreover, surprisingly, PAR% was higher in females (**Figure 2A**). Nevertheless, we found that female patients were older than males (**Figure 2B**) and PAR% showed a marginal positive correlation with age that correlates well with previous studies showing positive correlation between PARylation and age in humans [29-32] (**Figure 2B**). Importantly, PARylation indicies positiviely correlated with time of hospitalization (time to death) in males but not in females (**Figure 2C**).

**Figure 2.**
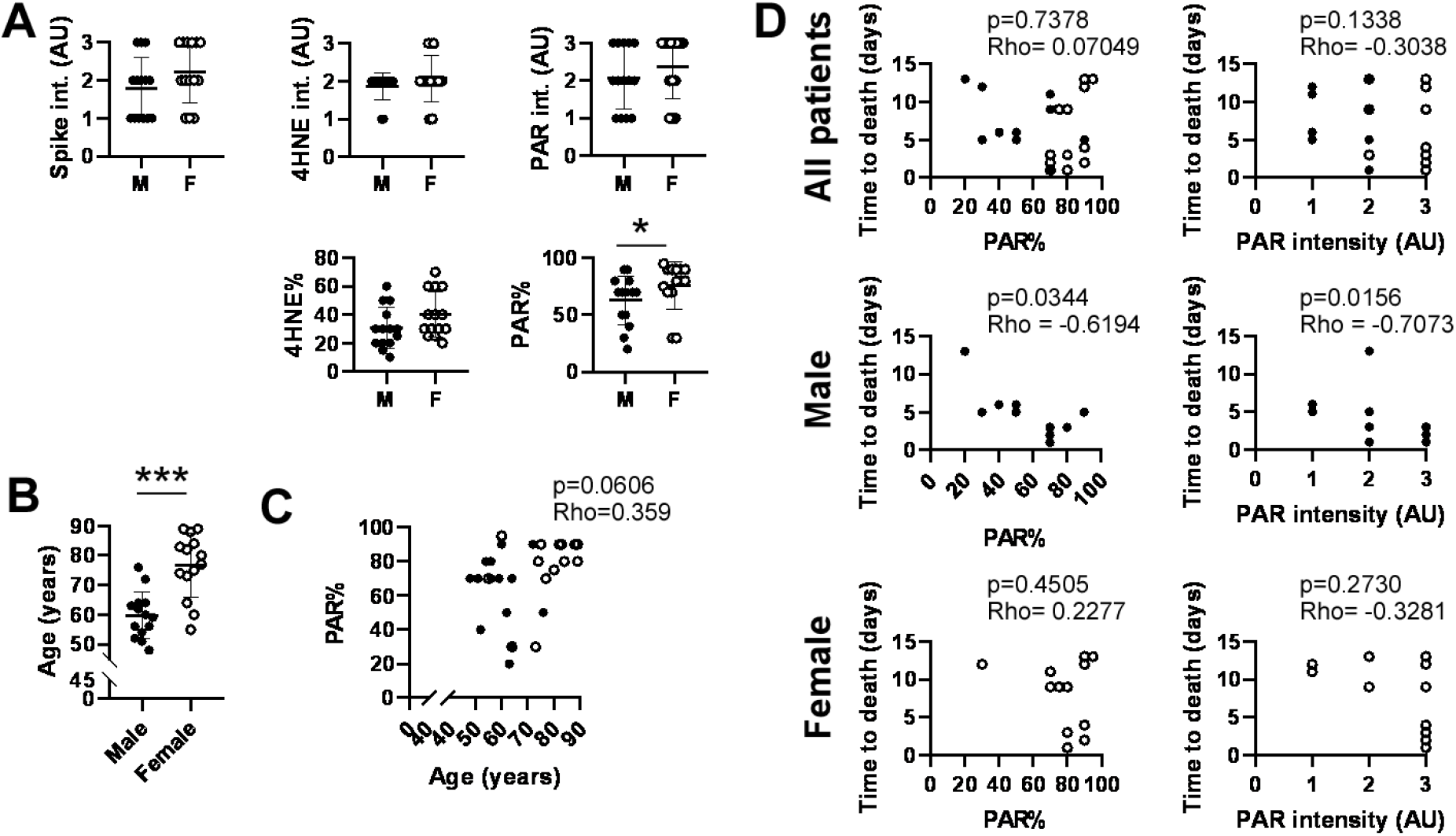
PARylation in the lungs negatively correlates with the time of hospitalization. **(A)**The indicated immunohistochemistry reactions were scored for the cases in the patient cohorts. Scale bar is 10 µm. Normality was checked using the Shapiro-Wilk test. In all cases Mann-Whitney test was applied to assess statistical significance. Average ± SD are depicted. **(B)**The age of the individuals was compared between males and females. Normality was checked using the Shapiro-Wilk test. Two-sided unpaired t-test was used to assess statistical significance. Average ± SD are depicted. **(C)**Correlation between age and PAR% was assessed using Spearman correlation. Normality was checked using the Shapiro-Wilk test. **(D)**Correlation between time to death and PARylation indices was assessed using Spearman correlation considering all patients, males and females. In two males hospitalization time was omitted due to their extended length. Normality was checked using the Shapiro-Wilk test. *, ** or *** symbolize statistically significant difference between the indicated groups at p<0.05, 0.01 or .0.001, respectively. Abbreviations: 4HNE – 4-hydroxynonenal, PAR – poly(ADP-ribose). Key: closed circles = male and open circles = female.

## Discussion

In this retrospective study pulmonary oxidative stress and PARylation was evidenced in COVID-19 patients similar to [2-7,56]. In COVID-19 patients oxidative stress and PARylation was present throughout the tissue. Oxidative stress, virus load and PARylation peaked in the exudative phase and decreased in the proliferative phase. Markers of oxidative stress and spike protein correlated with PARylation, however, there was only a borderline correlation between 4HNE indices (oxidative stress markers) and spike protein suggesting that oxidative stress and virus load are independent pathways, both converging on inducing PARylation. These observations are in line with our previous study [7] showing that spike protein or viral RNA are inducers of PARP-dependent inflammation.

When comparing males and females, we observed higher percent of PAR positive cells in females as compared to males. This observation contradicts to higher PARylation levels in males over females in non-COVID19 models [44-46]. However, there was a negative correlation between pulmonary PARylation and the time to death in males that accords with the higher fatality rate of COVID-19 among males [41-43,55] and with the higher sensitivity of males to PARylation-induced tissue damage [44-46]. In addition, male patients were younger than females and PAR levels increased with age in this cohort, similar to [29-32]. Apparently, males become vulnerable to PARylation-mediated tissue damage at an earlier age as compared to women. This may be related to the higher PARP activity reported in males compared with females under 60 years of age [44]. Furthermore, it is also possible that only women with high PARylation that get admitted to hospital. Taken together, PARylation can contribute to the higher male morality rate in COVID-19 [41-43,55] and the data also suggest that high PARylation levels may be an indication of worse disease outcome.

These data suggest that pulmonary oxidative stress and PARylation is involved in the pathogenesis of COVID-19. Oxidative stress and PARylation correlates with cell death and tissue damage (including kidneys and liver dysfunction), inflammation and blood clotting abnormalities (the patients in this study were screened for DIC), therefore, ultimately likely contribute to fatal outcomes [57,58].

Importantly, the retrospective nature of the study represents a limitation, due unknown co-morbidities, differences in the lifestyle choices and behavior between sexes and inconsistent timing of blood collection. Nevertheless, the results of the study build a case for repurposing of pharmacological PARP inhibitors for acute COVID-19. Through preventing early stage permanent tissue damage, early application of PARP inhibitors may preserve tissue structure and function that is likely to be beneficial in the subsequent post-COVID timeframe as well.

## Data Availability

Primary data associated to this manuscript can be found at https://figshare.com/s/725650d2036f586f065f (DOI: 10.6084/m9.figshare.22303120).

## Funding statement

Our work is supported by grants from the NKFIH (K141142, FK146852, TKP2021-EGA-19, TKP2021-EGA-20) and the Hungarian Academy of Sciences (POST-COVID2021-33, NKM2022-30). Project no. TKP2021-EGA-19 and TKP2021-EGA-20 has been implemented with the support provided from the National Research, Development and Innovation Fund of Hungary (NKFIH), financed under the TKP2021-EGA funding scheme. This project has received funding from the HUN-REN Hungarian Research Network. Supported by the University of Debrecen Program for Scientific Publication.

## Ethics approval statement

The study was authorized by the local ethical board (6043/2022).

## Patient consent statement

In the frame of the study post-mortem samples were assessed, hence patient consent is not needed.

## Permission to reproduce material from other sources

There is no such item in the manuscript.

## Clinical trial registration

Not applicable.

## Supplementary Description

The Supplementary Materials contain the 1) detailed description of immunohistochemistry, 2) the validation of the antibodies used and 3) a tabular collection of the statistical indices.

## Notes

**Conflict of interest disclosure:** The authors have no competing interest to disclose.

### Competing Interest Statement

The authors have declared no competing interest.

### Author Declarations

The study was authorized by the local ethical board of the University of Debrecen (6043/2022).

