## Supplementary materials for "Oxidative stress and PARP activation in the lungs is an early event in COVID-19 pneumonia"

### Supplementary methods

*Immunohistochemistry*

Immunohistochemistry was performed with anti-spike, anti-4-hydroxynonenal (4HNE) and anti-PAR antibodies. Five micron thick sections were obtained from formalin-fixed, paraffin embedded blocks of COVID-19 patients and controls. Tissues were formalin-fixed 24 hours after excision. Immunohistochemistry was performed as in [1] with the conditions described in **Supplementary Table S3**.

The specificity of the antibodies was assessed in the following way:

1) The anti-spike antibody gave signal only in the lung tissues of patients who had clinical signs of COVID-19 as tested hereby (**Supplementary Figure 1**) and in a previous study [2]. Changes to the virus load as a function of the disease stage, deducted from the staining using this antibody, correlates with the previous observations on virus load throughout the disease [3]. The antibody yields a cytosolic staining.
2) The PAR antibody (10H) is a validated, commonly accepted antibody directed against poly(ADP-ribose) polymers longer than 20 ADP-ribose units [4,5]. The antibody yields a nuclear staining.
3) 4HNE levels are good measures of oxidative stress [6]. The 4HNE antibody was validated to be responsive to oxidative stress in prior studies from us [7,8] and others [9,10]. Due to the pleiotropic, indiscriminate nature of oxidative stress, the antibody yields a diffuse staining throughout the tissue.

The overall intensity of staining intensity was scored between 0-3 (referred as an intensity value i.e. spike intensity, 4HNE intensity, PAR intensity) and the percent of positive cells (given as a percentage, i.e. 4HNE%, PAR%) was evaluated. Sample photographs representing intensity scores are on **Supplementary Figure 2**. All histology sections were reviewed and scored by two independent experts.

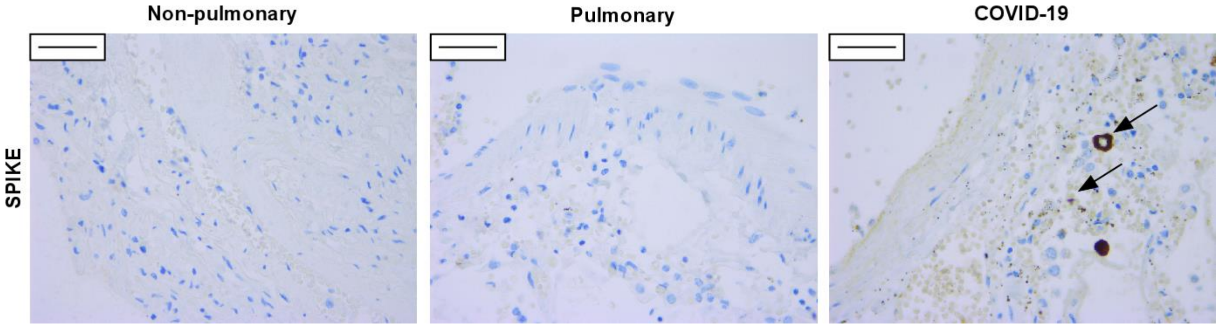

**Supplementary Figure 1. Positive spike staining is achieved only in the lung biopsies of patients with COVID-19.**

Immunohistochemistry was performed on the lung biopsies of patients died of non-pulmonary and pulmonary causes, as well as, due to COVID-19. Representative images are presented. Arrows point at positive cells. Scale bar is 50 μm.

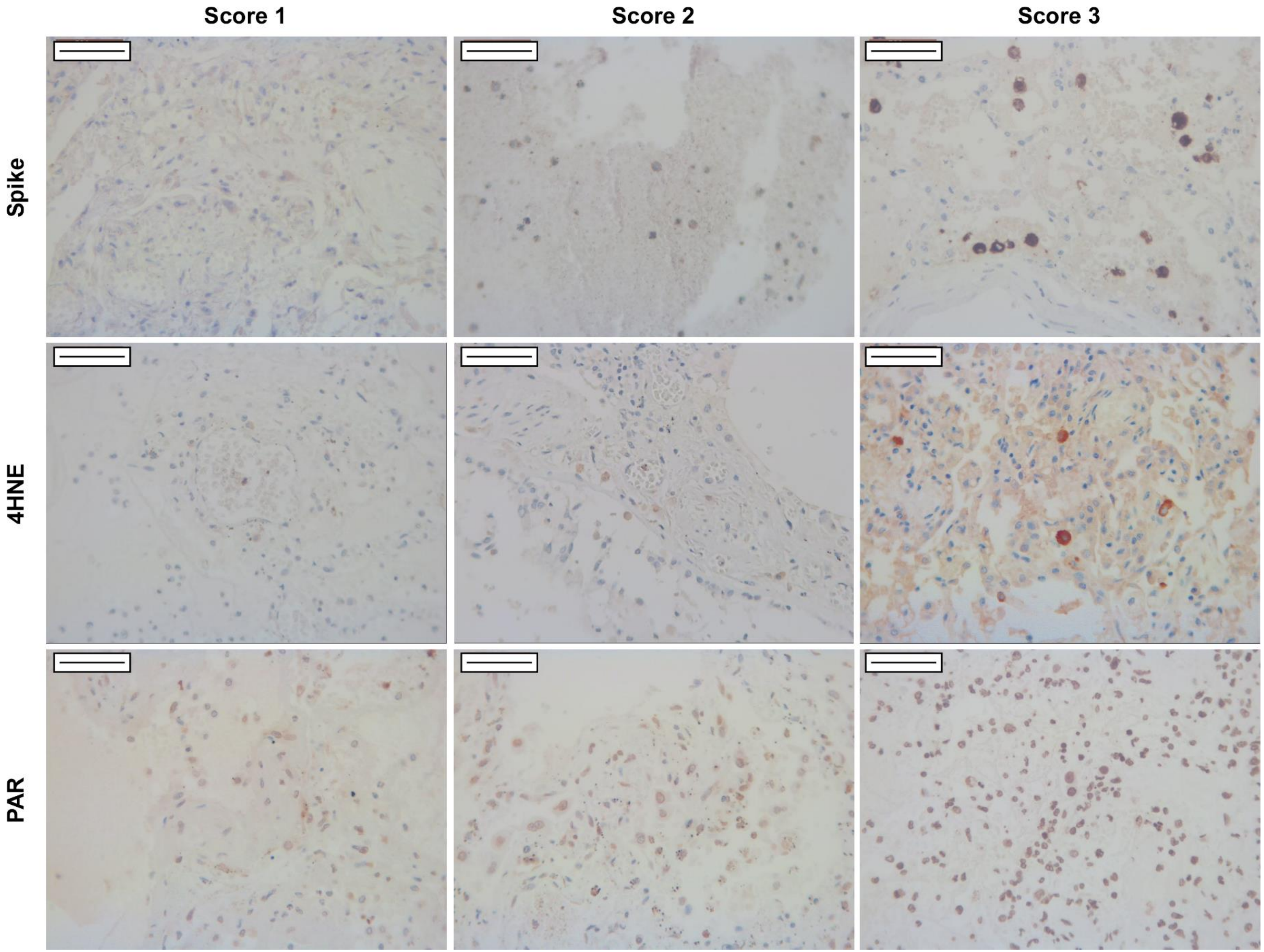
Score 1
Score 2
Score 3
Spike
4HNE
PAR

**Supplementary Figure 2. Sample coloration for each marker**

The bar is equivalent to 500 μm. Brightness was adjusted to level intensities.

**Supplementary Tables**

**Supplementary Table 1. Conditions for immunohistochemistry**

Abbreviation: CC1 - ULTRA Cell Conditioning Solution (Ventana Medical Systems, Oro Valley, AZ, USA)

| Primary Antibody | | | | | Antigen retrieval: HIER | | Immunohistochemical stainers | | | |
|---|---|---|---|---|---|---|---|---|---|---|
| **Antibody name** | **Vendor** | **Host** | **Dilution factor** | **Incubation time/temp.** | **Method** | **Time/Temp.** | **Visualization system** | **Enhancement** | **Counterstain** | **Staining Platform** |
| **Anti-4 Hydroxynonenal antibody (ab46545)** | **Abcam** | rabbit polyclonal | 1/400 | 32'/37 °C | CC1, pH8,5 | 36' ',95°C; | UltraView Universal DAB Detection Kit | $CuSO_4$ | Hematoxylin II | VENTANA BenchMark ULTRA |
| **Poly(ADP-ribose) monoclonal antibody (10H). antibody (10H). (ALX-804-220-R100)** | **Enzo** | mouse monoclonal | 1/2000 | 32'/37 °C | | 64 ',95°C; | | | | |
| **SARS-CoV-2 spike** | **Abcam** | rabbit polyclonal | 1/1000 | 1h /37 °C | | 56 ',100°C; | | | | |

| |
|---|
| **Glycoprotein S1 ab/ (ab275759)** |

**Supplementary Table 2. Correlation coefficients between the histological indices in patient groups**

Normality was checked using the Shapiro-Wilk test then Pearson correlation was calculated for variables with normal distribution and Spearman correlation was calculated for variables with non-normal distribution. *, ** or *** symbolize statistically significant difference between the indicated groups at p<0.05, 0.01 or .0.001, respectively.

| Patient group | Dataset | | p | Rho | Test |
|---|---|---|---|---|---|
| | Variable 1 | Variable 2 | | | |
| All cases | spike intensity | 4HNE intensity | 0.06 | 0.360 | Spearman |
| | | 4HNE% | 0.117 | 0.303 | Spearman |
| | | **PAR intensity** | **0.003 **** | **0.561** | **Spearman** |
| | | **PAR%** | **0.0001579 ***** | **0.664** | **Spearman** |
| | 4HNE intensity | **PAR intensity** | **0.000455 ***** | **0.618** | **Spearman** |
| | | **PAR%** | **0.001 ***** | **0.614** | **Pearson** |
| | 4HNE% | PAR intensity | 0.366 | 0.178 | Spearman |
| | | PAR% | 0.085 | 0.331 | Pearson |
| Exudative | spike intensity | 4HNE intensity | 0.959 | -0.088 | Spearman |
| | | 4HNE% | 0.148 | 0.366 | Pearson |
| | | PAR intensity | 0.567 | 0.150 | Pearson |
| | | **PAR%** | **0.031 *** | **0.906** | **Pearson** |
| | 4HNE intensity | PAR intensity | 0.541 | 0.257 | Spearman |
| | | **PAR%** | **0.037 *** | **0.558** | **Spearman** |
| | 4HNE% | PAR intensity | 0.531 | 0.163 | Pearson |
| | | PAR% | 0.224 | 0.311 | Pearson |
| Proliferative | spike intensity | 4HNE intensity | 0.076 | 0.830 | Spearman |
| | | 4HNE% | 0.220 | 0.407 | Spearman |
| | | PAR intensity | 1.000 | 0.038 | Spearman |
| | | PAR% | 0.055 | 0.593 | Spearman |

| | | | | | |
|---|---|---|---|---|---|
| | 4HNE intensity | PAR intensity | 0.194 | 0.571 | Spearman |
| | | PAR% | 0.367 | 0.305 | Spearman |
| | 4HNE% | PAR intensity | 0.476 | 0.275 | Spearman |
| | | PAR% | 0.052 | 0.597 | Pearson |
